# Did national holidays accelerate COVID-19 diffusion in Bangladesh? A case study

**DOI:** 10.1101/2022.03.16.22272515

**Authors:** Mohammad Radid Khan, Sayem Mottakin, Md. Ahsan Kabir, Md. Enamul Hoque, Mohammad Ruhul Amin, Md. Shariful Islam

**Affiliations:** Department of Biochemistry and Microbiology, North South University, Dhaka, Bangladesh; Department of Electrical and Computer Engineering, North South University, Dhaka, Bangladesh; Department of Physics, Shahjalal University of Science and Technology, Sylhet, Bangladesh; Computer and Information Science, Fordham University, New York, USA; Department of Mathematics and Physics, North South University, Dhaka, Bangladesh

**Author notes:** Corresponding author: Md. Shariful Islam.

**Keywords:** COVID-19, SARS-CoV-2, National events, R_t_ values, Bangladesh

## Abstract

Bangladesh registered 1573828 confirmed cases of COVID-19 and the death toll crossed the grim milestone of 27946 across the country as of 9th December, 2021. Despite the enforcement of stringent COVID-19 measures, including nationwide lockdowns, travel bans, tighter curbs on nonessential activities, and social distancing, the country witnessed an accelerated diffusion of coronavirus cases during the national events and festivals in 2020. The present study aims to examine the association between the national events / festivals and the transmission dynamics of COVID-19 by looking at the instantaneous reproduction number, R_t_, of the 64 districts in Bangladesh. We further illustrate the COVID-19 diffusion explicitly in Dhaka Division at the first phase of the pandemic. The comprehensive analysis shows an escalation of R_t_ value in Dhaka and in all industrialized cities during the major events such as, Garments reopening and religious holidays in Bangladesh. Based on the analysis, a set of array measurements has been also suggested to evade the future pandemic risks while running the national festival activities.

**Highlights:**

- Bangladesh confirmed 1573828 coronavirus cases and 27946 deaths due to the current COVID-19 outbreak.
- Country observed significant COVID-19 diffusion in its business hubs during national holidays.
- Dhaka, the capital of Bangladesh, is the epicenter of the ongoing pandemic.
- Calculated R_t_ value illustrates its escalation in Dhaka and its neighboring cities at the time of national events.
- Bangladesh Government needs to consider interdisciplinary approaches and contextual policies to contain the future pandemic during any national events.

## Introduction

Ever since the end of 2019, the world has been afflicted by coronavirus disease 19 (Covid-19), which is caused by the Severe Acute Respiratory Syndrome Coronavirus (SARS-CoV-2019) [1]. This deadly virus has wreaked havoc upon the global population, and has already taken the lives of 5.18 million people as of late November, 2021 [2]. As COVID-19 has infectious characteristics [1], it rapidly spread out around the world, with the United States of America currently leading the weekly death charts, followed by the Russian Federation and Ukraine [3]. While Wuhan-1 D614G lineage was the primary variant of SARS-CoV-2019 at the early stage of this global outbreak, multiple variants have emerged over time, often with individual countries having unique dominant strains [4]. Soon after the Bangladeshi capital city of Dhaka began experiencing the first wave of COVID-19 in early March 2020, the WHO formally termed COVID-19 to be a pandemic. It wasn’t long before the local government announced the first lockdown near the end of March 2020, to mitigate the diffusion of the virus [5,6]. The stringent lockdown played a pivotal role in reducing the COVID-19 transmission at the initial stage of the pandemic, with the criticality of this restriction becoming quickly apparent, considering the current socio-economic status of Bangladesh [7]. Therefore, the government seemingly prioritized the economy, during the earlier stages of the pandemic, and evidently decided to adopt a flexible lockdown system with the gradual opening of all industries and sectors of commerce, apart from educational institutions [8]. Since Dhaka, the capital of Bangladesh, one of the most crowded cities in the world, carries up to 35% of the country’s economy [8], it is not surprising why Dhaka became the epicenter of the current pandemic. Besides this metropolitan city, its surrounding areas including Gazipur and Narayanganj also hold financial significance as important industrial zones. Multiple lines of evidence also reported the high diffusion of coronavirus in these domains in 2020 [9].

An epidemic will occur if R_t_ value, instantaneous reproduction number, is greater than 1, whereas the number of cases will drop when R_t_ < 1[10]. The R_t_ values provide quantitative real-time feedback that represents how the population being observed is faring under present conditions. As such, varying the conditions through various means, often government-imposed regulations such as lockdowns, having to wear masks when outdoors, and so on, will allow to see trends in the R_t_ values that show the effectiveness of tactics in place. The aim of this study is to explore the evolution of the coronavirus infection cases in Dhaka and its neighboring cities at the first phase of the pandemic and examine the impact of national events on the pandemic by looking at the R_t_, over time. We notice the spontaneous trend of COVID-19 cases particularly in Dhaka division regardless of the pandemic situation in other districts, while observing the mass movement due to the major national events in Bangladesh. Notably, the R_t_ value goes over 1 on multiple occasions in Dhaka and a few other cities during major events or festivals such as garments factories reopening time, Eid Ul-Fitr and Eid Ul-Adha (religious holidays for Muslims) in 2020.

## Methods

The basic reproduction number (R_0_) defines the transmissibility of the disease. It is the mean of the variable number of secondary infections brought about by the primary infection, in a totally susceptible population [10]. When R_0_ > 1, an epidemic will occur within the population. However, realistically, populations will rarely be totally susceptible, for a myriad of reasons [11]. The effective reproduction number (R_e_) accounts for this assumption made by the basic reproduction number, and calculates an adjusted value. When it is calculated in real-time, for time “t”, it is referred to as R_t_. When R_t_ > 1, an epidemic will occur. If R_t_ = 1, then the number of cases is stable, whereas the number of cases will start dropping when R_t_ < 1 and the disease eventually ceases to be an epidemic or endemic.

This study calculates the R_t_ values for the 64 districts of Bangladesh including the Dhaka and Chattogram Division, and analyses the results to identify the trends with respect to various major events and festivals. The R_0_, and R_e_ values are calculated by the following formulas:

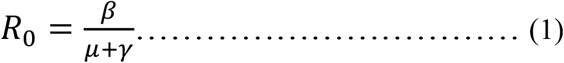

Where β, *γ, μ* represents infection rate, death rate, and recovery rate respectively.

R_e_ can be written as,

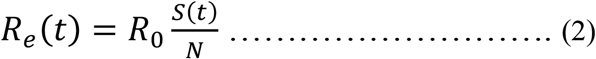

From the SIRD model [6], we know that,

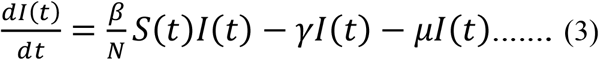

With the help of Eq. (3), when *I = I*_*max*_, it can be derived that *S(t) = N / R*_*0*_ and *R*_*e*_ *= 1*.

Here, “t”, “S”, and “N” denote time “t”, susceptible population, and total population respectively, whereas random walk is represented by θ, and *γ*′ varies independently [6].

In order to calculate R_t_, we establish a link between the initial number of confirmed cases (*y*_*j*_) and the number of cases of a previous day, right before a specific *T*_*s*_. We can equate this idea as shown below:

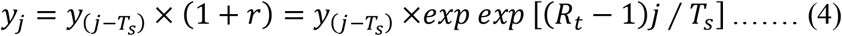

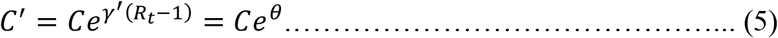

Here, *γ*′varies independently, while random walk is observed by C = y_j_ and *ϑ*=*γ*′(*R*_*t*_ − 1). The term “serial interval” (*T*_*s*_) refers to the length of time that passes between subsequent cases in a series of transmissions of a particular disease.

Thus, solving the above equations, *R*_*t*_ can be written as

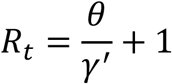

It is important to be able to calculate the instantaneous reproduction number, since monitoring its value and comparing it with the ratio between the total population and the susceptible population in the early stages will tell us when the ensuing pandemic goes beyond our control [6].

### Data visualization

This study uses Python, Pandas, GeoPandas, Spyder IDE and ArcMap to build the map. Python is embedded into Geographical Information System (GIS) applications for this study. ArcMap is applied to create, edit and view geospatial data. GeoPandas is a Python module based on the Pandas library that makes it easier to work with spatial data. We have taken the spatial map of Bangladesh from HDX (The Humanitarian Data Exchange) and separated the 13 districts of the Dhaka division from the downloaded shapefile using ArcMAP.

Here, each District is represented in the form of a single polygon in a shape file and it was converted into a Geopandas GeoDataFrame. We converted the data collected from the CSV file into a Pandas DataFrame and performed a join operation in python to combine both the GeoDataFrame and DataFrame into one GeoDataFrame. Lastly, by using the GeoDataFrame, we visualized the map using ArcMAP software.

R_t_ values for different districts were calculated by using Python and the collected data were visualized in R, with RStudio as the IDE. With the help of the package “ggplot2”, graphs of R_t_ vs time were plotted for all 64 districts of Bangladesh. Finally, with the help of the “cowplot” package, the individual graphs were all combined into multi-plot graphs.

## Results

### Diffusion of COVID-19 at the first phase of pandemic in Dhaka division

To explore the diffusion of coronavirus, first we checked the number of infected cases in Dhaka division over time as this zone carries major economic significance. At the beginning of March 2020, only 2 out of the 13 districts, that is, the capital Dhaka and the neighboring Narayanganj, were infected with Covid-19 as shown in Fig. 1. However, the disease spread quickly within the Division in the oncoming months due to Dhaka district being the primary commercial hub of the region, and thus deeply economically connected to the bordering districts. During April and May, rapid transmission occurred within the northern Districts such as Gazipur and Narsingdi, only to then disseminate much more swiftly throughout the eastern and southern ones, namely Manikganj, Rajbari, Faridpur, Gopalganj, Madaripur, and Shariatpur, in June, as reflected by the number of cases per 10,000 (Fig.1). Eventually, the total number of new cases throughout Dhaka Division peaked around the first week of July at ∼ 1050 cases. The overall situation in the various Districts during August 2020 remained similar to July. Notably, it was only in September 2020 that the conditions improved in the Division, primarily in the eastern and southern districts such as Faridpur, Gopalganj and Shariatpur, with the gradual improvements making their way into the other districts, namely Manikganj, Rajbari, and Madaripur, during October 2020 as well. We speculate that the enforcement of several stringent measures including wearing masks, maintaining social distance and extending the lockdown by 66 days are associated with the strongest and most widespread attenuation of COVID-19 cases in Bangladesh. However, even in October, the state of affairs in Dhaka district remained at an alarming level (Fig. 1). To understand more clearly how the disease spread throughout the division, next we aim to examine the dynamics of R_t_ values during this period in Dhaka division.

**Fig 1.**
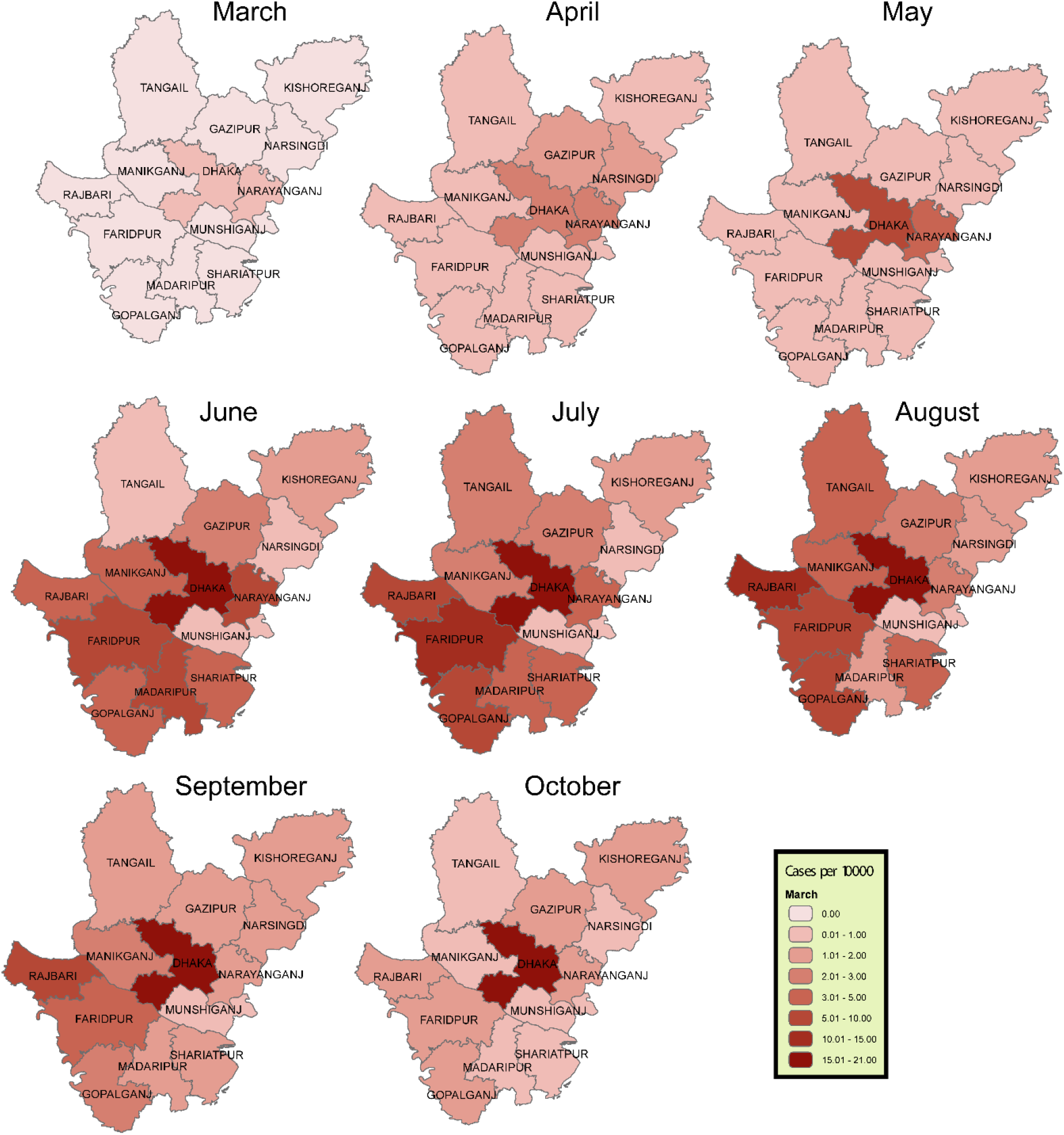
Propagation of COVID-19 from March, 2020 to October, 2020 in Dhaka Division

### R_t_ dynamics of COVID-19 in Dhaka Division during national events

Bangladesh has witnessed the occurrence of new local outbreaks during the national holidays in 2020. We hypothesize that these outbreaks are associated with the increased inter-regional mobility and thus increase the effective reproduction number (R_t_). To examine the evolution of the R_t_ values in Dhaka Division, a segmented regression analysis has been performed for the first 8 months (March 2020-October 2020) of the pandemic. While the R_t_ value for Dhaka and Gazipur districts stayed above 1 for the majority of the duration (8 months) covered by our data, the same cannot be said for the rest of the Division (Fig. 2). Apart from these two, all other districts maintained an R_t_ value of less than 1 throughout the time period being investigated, with peaks and troughs spread out primarily around major events, such as when garments reopened (Major event 2), Eid-ul-fitr (Major Religious Festival 1) and Eid-ul-Adha (Major Religious Festival 2). Mass-testing with RT-PCR kits began a couple weeks after the first lockdown (Major Event 1), in Dhaka [12], followed by other districts [13,14], which is reflected by the spikes in the R_t_ values for these districts in mid-April or soon after. Similarly, since testing began at different times in the rest of the districts, they show spikes in their values much later, in the following months. Dhaka, Gazipur, and Narayanganj comprise the primary locations of the garments factories in the Division. Initially, before Major Event 1(ME1), none of the districts had any R_t_ values because of unavailable data. However, after the ME1, some districts experienced a large spike in R_t_ values, with the way being led by Dhaka, Gazipur and Narayanganj (Fig. 2). The R_t_ value for Dhaka rapidly decreased leading up to Major Event 2(ME2), only to increase again afterwards. For Gazipur however, the R_t_ values increased leading up to ME2 followed by an attenuation trend afterwards. On the other hand, the R_t_ values started decreasing right before ME2, and kept doing so afterwards in Narayanganj. These spontaneous characteristics of R_t_ values reflect the COVID-19 transmission as a result of the mass workers moving throughout the districts to get to their workplaces as soon as they opened.

**Fig 2.**
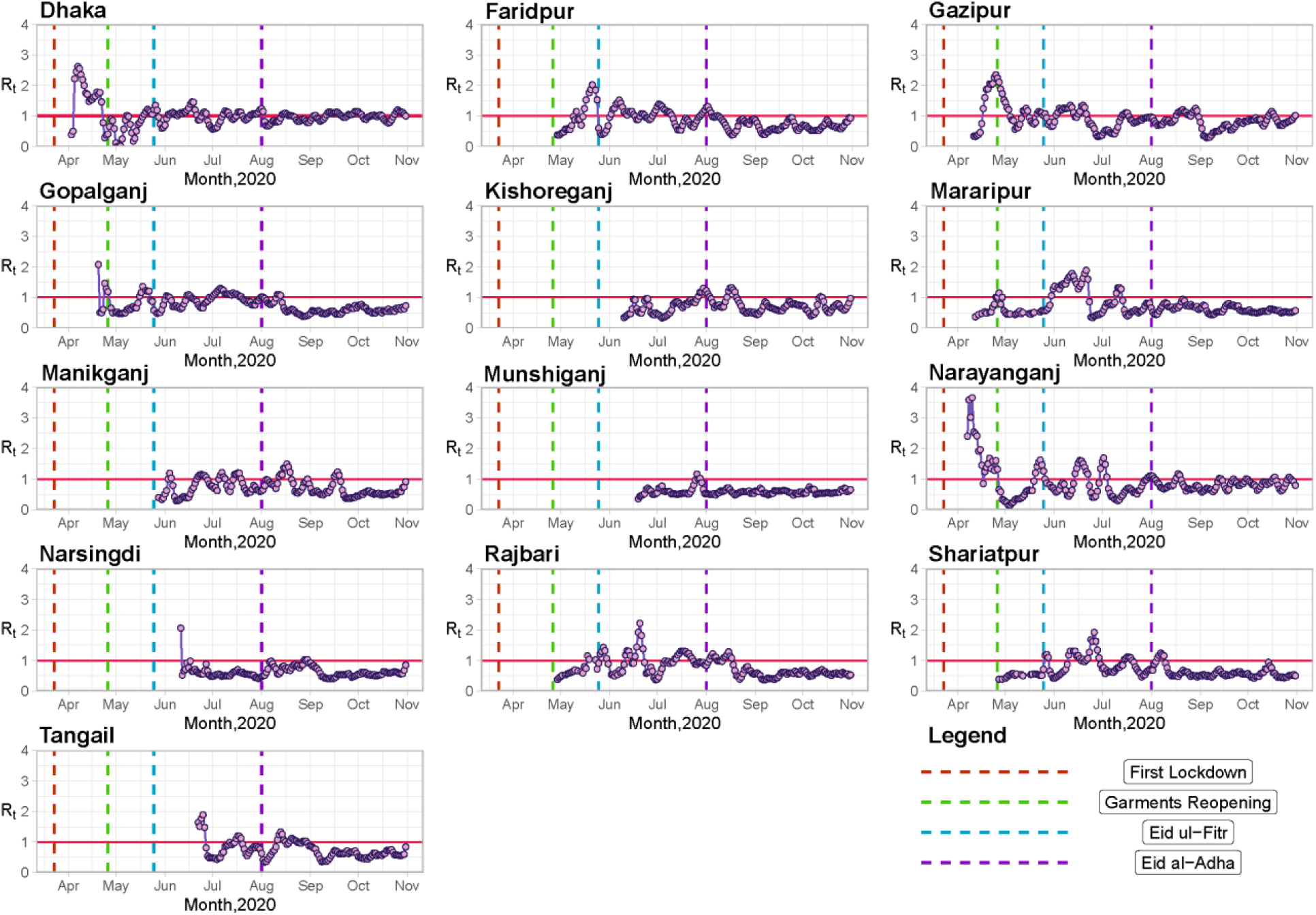
Comprehensive analysis of R_t_ values over time (March, 2020 – October, 2020) in Dhaka Division

Now, when it comes to Major Religious Festival 1(MRF1), Dhaka had a slight rise, followed by a significant drop, and then a large increase in R_t_ values (Fig. 2). The decrease brought it to below 1, while the final increase left it much higher than 1. It was very similar for every other district with varying degrees of decreases and increases with a notable exception of Faridpur, showing a surge of R_t_ in the moments before the MRF1. Apart from these, few other districts such as Munshiganj, Kishoreganj, Tangail and Narsingdi did not have any R_t_ data at this time period. For Major Religious Festival 2 (MRF2), every single District had an increase in R_t_ values until just before, or right after the day of MRF2. Notably, we have also noticed a significant fall off followed by a moderate or sharp elevation of R_t_ that brought the majority of the districts above 1 again, such as Dhaka, Gazipur and Kishoreganj. However, the escalation was temporary for most districts as they soon returned to below 1. Even Kishoreganj, which had a surge, experienced a rapid attenuation of R_t_ values. This trend might speculate that people moved to their hometowns to celebrate the festivals with their families, and thus, resulted in a spike in the R_t_ values during such events.

#### Evolution of R_t_ values in Chattogram Division at the time of national events

Next, we check the trend of R_t_ values in Chattogram division during the national events. To do that, another R_t_ multiplot has been made by using the data from the 11 districts of Chattogram division. One of these 11 districts, Bandarban, did not have any available data throughout the time period investigated in this paper. Both Cox’s Bazar and Chattogram districts experienced significant spikes in their R_t_ values (Fig. 3). The rest of the districts manifested similar trends, with varying fluctuations. Strikingly, we also observed a spontaneous trend of R_t_ values in other major cities including Mymensingh, Sylhet, Pabna and Jaipurhat in the course of Garments reopening events (Suppl. Figs. 1-3). Moreover, the comprehensive analysis illustrates a surge of R_t_ value (∼1.8) in Barisal district during the MRF1 (S4. Fig).

**Fig 3.**
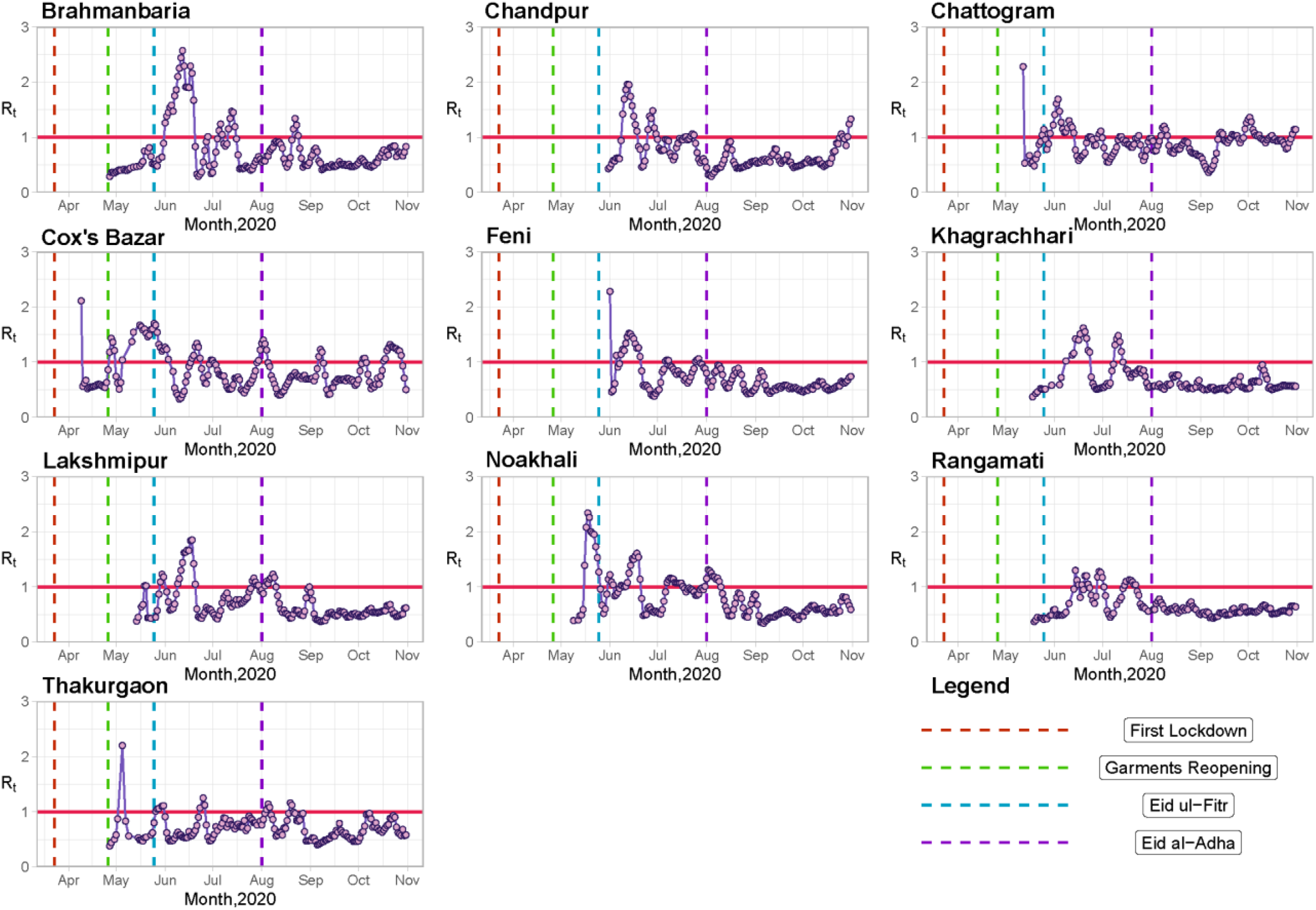
Dynamics of R_t_ values over time (March, 2020 – October, 2020) in Chattogram Division

## Discussion and Conclusion

While the Covid-19 pandemic has shaken all the countries throughout the globe to their very roots, the responses of individual nations to such a novel threat to humanity has varied greatly. Even when we look at the countries in the South Asian region, including Bangladesh, measures taken against the pandemic have differed significantly. This study looks at the strategies implemented by the Bangladeshi government in Dhaka Division during the national holidays or major festivals, and takes assistance from multiple statistics, illustrating them, analyzing the effectiveness of the measures taken and how they can be improved upon. Although the initial cases were reported in early March 2020, with a gradual increase in the number of daily cases, it was only from early May 2020 onwards that the number of daily infections began to rise exponentially. While the numbers represented the entire country, it was the commercial hubs of the nation that suffered the most, both economically and rate-of-infection-wise, as foreseen by Islam et al. [15]. The primary industry in the division is the RMG sector, operating mostly from Dhaka, Narayanganj, and Gazipur. As a result of the high demand for workers in RMG factories, the majority of employees originate from other districts, having moved to these industrial zones for work [16]. Due to the dense population of the division, with Dhaka city being one of the most densely populated cities in the world, it was not surprising when the government officials decided to impose the first lockdown in late March 2020 [5,6]. However, one of the negative impacts of the lockdown was that, since all workplaces, including the factories of the RMG sector, closed to abide by lockdown rules and regulations, most of these out-of-district workers, their families, or both, relocated back to their hometowns [17]. As a result, such high mobility of individuals led to greater risks of transmitting the virus. This explains the spikes in R_t_ values for Dhaka, Gazipur, and Narayanganj right after the first lockdown. Similar trends were observed when there was similarly high mobility among the masses during the holidays accompanying Eid ul-Fitr and Eid al-Adha [16,18–20]. Moreover, Dhaka is the major financial hub of the country and all the major commercial trade routes go through it. Hence, there is a constant influx of masses of people and transport both into and out of the district all the time. Due to its high physical communication with the rest of the districts, as well as the country, transmissibility of the virus is greatly heightened. This further leads to increases in the R_t_ of Dhaka district, as well as the rest of the districts. Chattogram district and Cox’s Bazar district are both important economic zones in the division, with the former being the primary port city in the country, and the latter being a world renown tourist hotspot. As such, both locations observed a rapid increase in the mobility of the population to and from the districts right before and after the major events and major religious festivals, be it for tourism or for the gradual revival of various other industries [21,22]. This resulted in both districts showing spikes in their R_t_ plots during these times. The spikes (R_t_ value ∼ 1.6) are especially prominent for Cox’s Bazar as the major events and religious festivals are often accompanied by somewhat lengthy national holidays, which further encourage mobility among the masses.

Despite the first cases of covid-19 being detected in the division on 8th March 2020, the first lockdown was only imposed on 23rd March 2020. Therefore, if the lockdown were imposed sooner, the transmission of the disease throughout the country could have been minimized as reported previously [8]. Recently, India has also been experiencing a sudden rise in COVID-19 transmission because of minimal lockdowns being imposed by the Indian government during the first half of 2021, while allowing celebration of religious festivals, alongside political rallies, with effectively no hindrance [23]. In both countries, unrestricted mobility of the general population, including asymptomatic victims of the virus, lead to the disease spreading rapidly through the respective populations. Therefore, an evidence-based approach and contextual strategies should be adopted to avoid the COVID-19 diffusion during future festivals. On the other hand, despite the numbers of experts constantly attempting to alert the people about the risks of covid-19, in order to help them understand the dire circumstances we find ourselves in, a significant portion of the masses refuse to even try to comprehend the present state of affairs, instead opting to publicly and aggressively demand their former unrestrained mobility, ignoring the directives issued by the government [7]. In addition to the Bangladeshi Government’s prescribed current cautionary measures, we suggest taking the following interdisciplinary approaches and contextual policies to contain future pandemics while running the national festival activities, further building on those recommended by [6,15]. Firstly, the closure and isolation of the major commercial hubs, such as Dhaka district, is crucial to limit the mobility of the general public and thus, minimize the spread of the disease to the neighboring districts. Secondly, government holidays for the national festivals must be kept at the bare minimum, since it has been observed that even holidays of moderate lengths lead to people going back to their hometowns, often carrying the virus with them. Similarly, a person might get infected when they visit their hometown and bring back the disease to their workplace after the holidays, further propagating the spread of the virus in the commercial zones. Thirdly, greater efforts must be made to raise awareness among the general populace in order to reduce misconceptions about the current situation, and make them more willing to take the necessary precautions to minimize the transmission of covid-19. Above all, we need to understand that this might be the beginning of a new humanitarian crisis and we might experience many more in the future. Therefore, if we do not upgrade ourselves to implement the best practices to save lives and do not learn the lessons from 2020, Bangladesh might experience another catastrophic situation in the near future.

## Data Availability

All data (case number) were collected from the IEDCR, Bangladesh website.

## Acknowledgement

We acknowledge the contribution of www.Pipilika.com software development team for collecting the daily data of total COVID-19 tests, total positive cases, and total deaths in Bangladesh. The authors would like to thank Dr. Fariha Jasmin Chowdhury for her critical comments as a general reader.

## Author’s contributions

MRK, SM, MAK, MI made substantial contributions to the conception or design of the work. MRK, SM and MAK completed the relevant studies and created plots related to the R_t_ values, while MRK and SM wrote the draft. MH, MA and MI critically revised the manuscript. All the authors met regularly to discuss the outcome of each experiment. The data were collected from the IEDCR website and each of the results was cross checked by at least three of the six authors. All authors are responsible for acquiring, analyzing, and interpreting the data for this article.

## Availability of data and materials

All data were collected from the IEDCR, Bangladesh website.

## Ethics approval and consent to participate

Not applicable

## Consent for publication

Not applicable

---

**S1 Fig.**
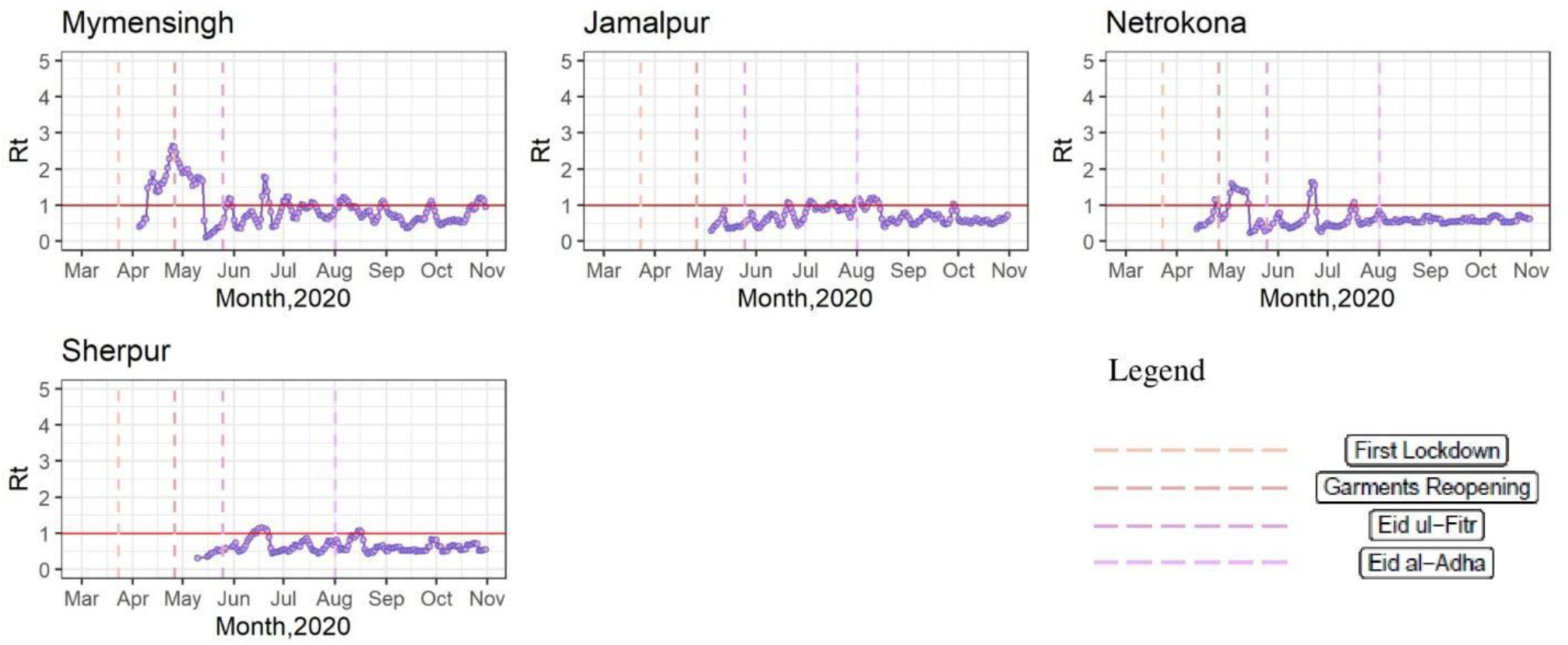
Dynamics of R_t_ values over time (March, 2020 – October, 2020) in Mymensingh Division

**S2 Fig:**
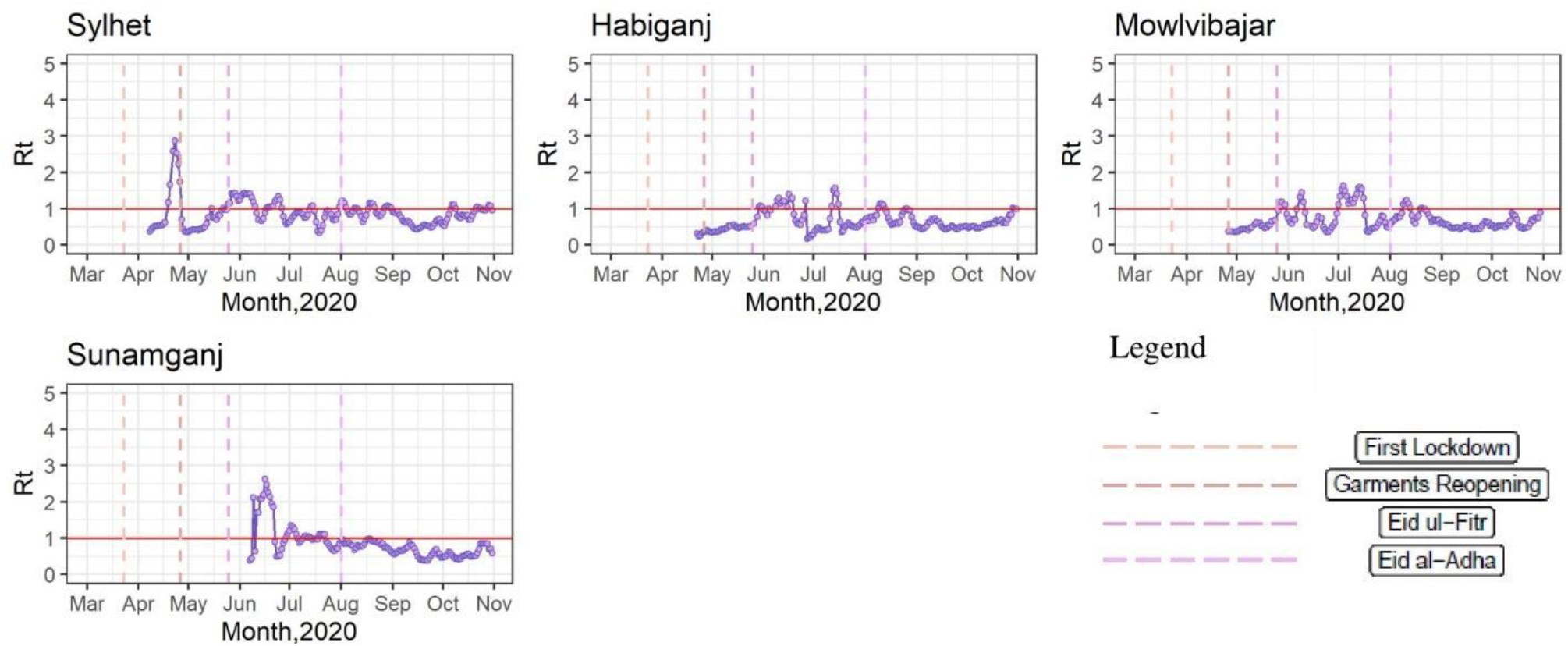
Dynamics of R_t_ values over time (March, 2020 – October, 2020) in Sylhet Division

**S3 Fig:**
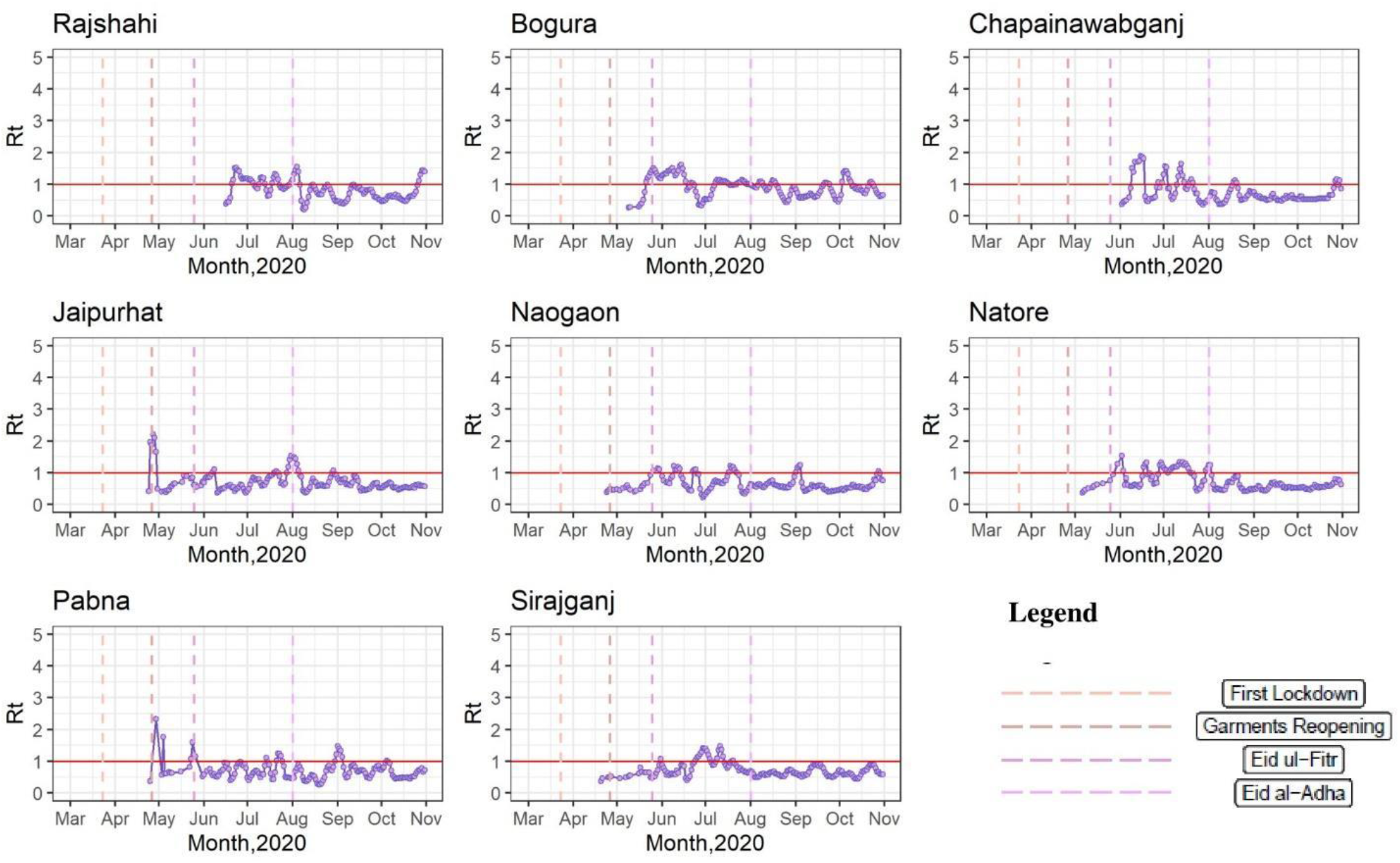
Dynamics of R_t_ values over time (March, 2020 – October, 2020) in Rajshahi Division

**S4 Fig:**
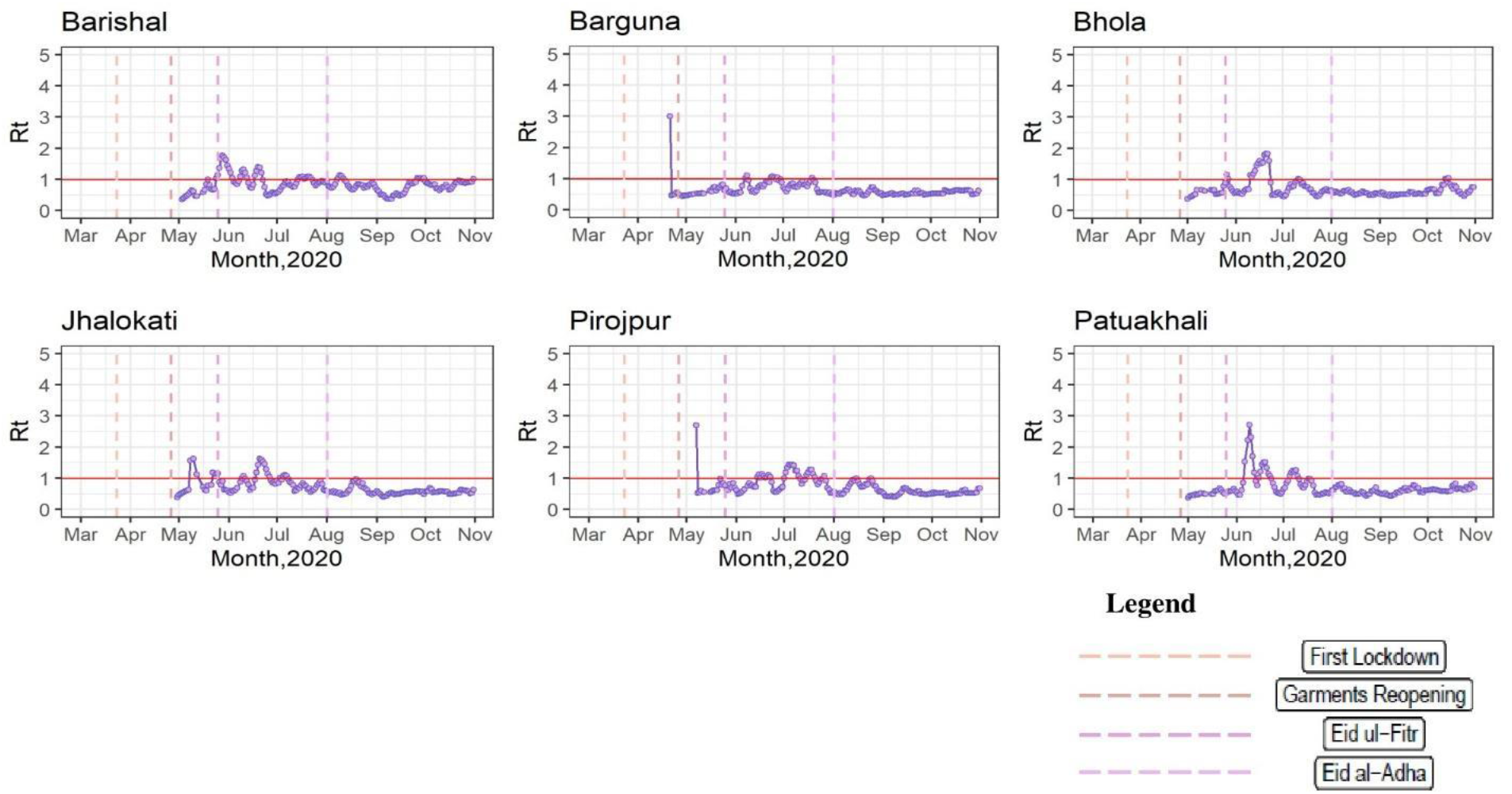
Dynamics of R_t_ values over time (March, 2020 – October, 2020) in Barisal Division

**S5 Fig:**
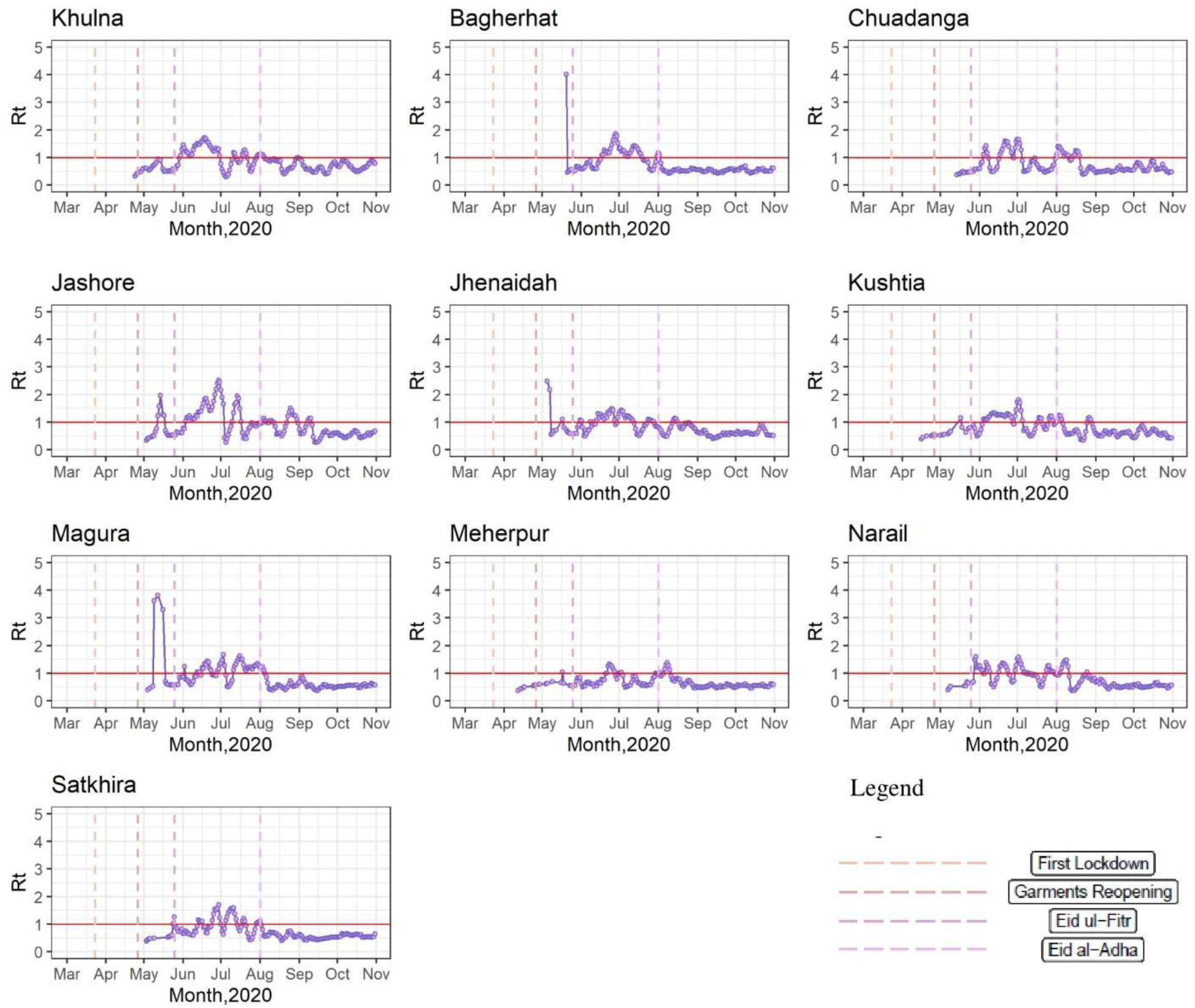
Dynamics of R_t_ values over time (March, 2020 – October, 2020) in Khulna Division

**S6 Fig:**
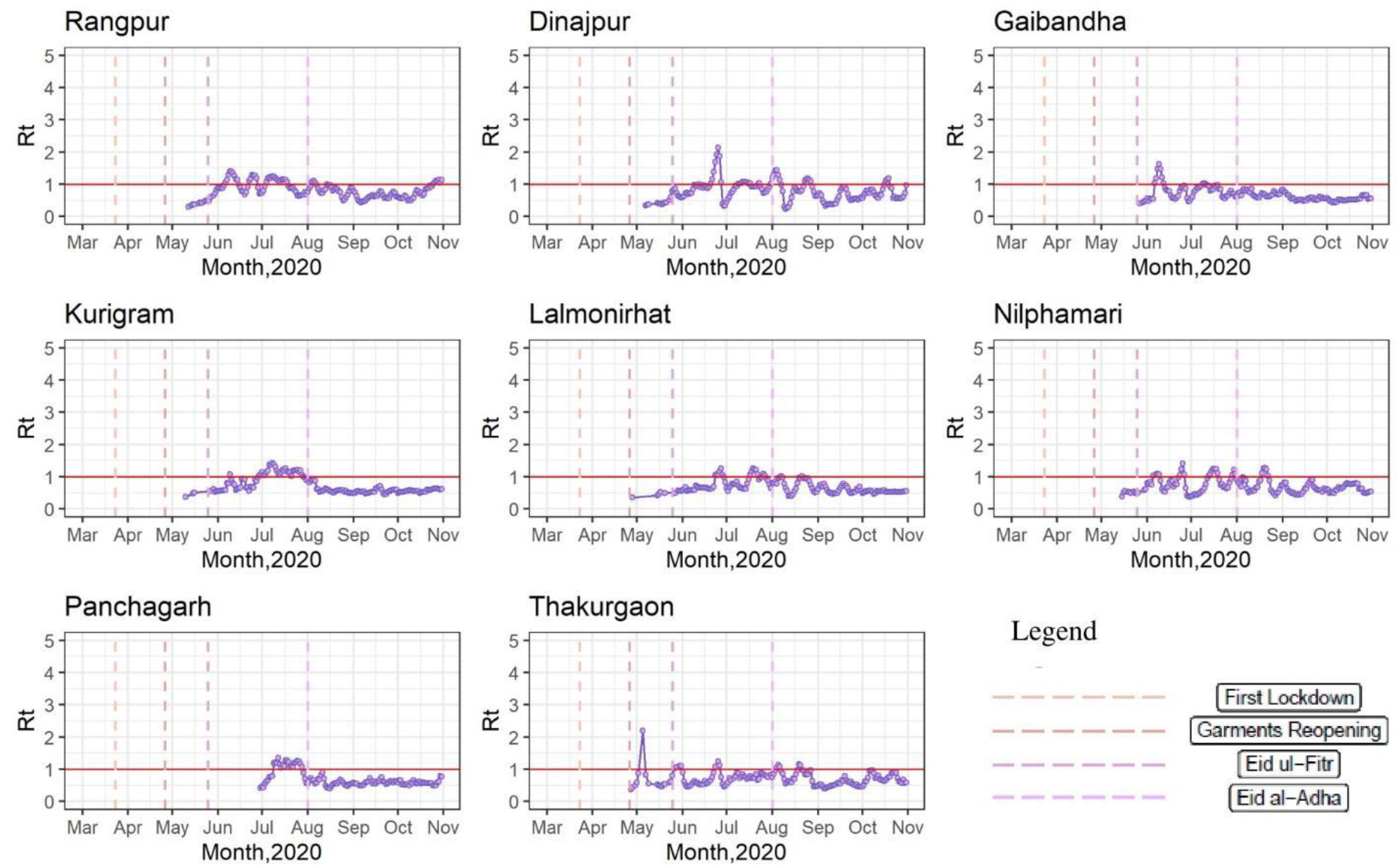
Dynamics of R_t_ values over time (March, 2020 – October, 2020) in Rangpur Division

